# Automatic speaker diarization for natural conversation analysis in autism clinical trials

**DOI:** 10.1101/2023.05.31.23290782

**Authors:** James O’Sullivan, Guy Bogaarts, Philipp Schoenenberger, Julian Tillmann, David Slater, Nima Mesgarani, Eckhart Eule, Timothy Kilchenmann, Lorraine Murtagh, Joerg Hipp, Michael Lindemann, Florian Lipsmeier, Wei-Yi Cheng, David Nobbs, Christopher Chatham

**Affiliations:** Roche Innovation Center Basel, F. Hoffmann-La Roche Ltd., Basel, Switzerland; Biomarkers & Translational Technology, Neuroscience & Rare Diseases, Pharma Research & Early Development, Roche Innovation Center NY; Roche Innovation Center New York, Roche TCRC Inc., New York, USA; Neuroscience Early Development, Pharma Research & Early Development, Roche Innovation Center Basel, Basel, Switzerland; Neuroscience & Rare Diseases, Pharma Research & Early Development, Roche Innovation Center Basel, Basel, Switzerland; Mortimer B. Zuckerman Mind Brain Behavior Institute, Columbia University, New York, NY 10027, United States

## Abstract

Challenges in social communication is one of the core symptom domains in autism spectrum disorder (ASD). Novel therapies are under development to help individuals with these challenges, however the ability to show a benefit is dependent on a sensitive and reliable measure of treatment effect. Currently, measuring these deficits requires the use of time-consuming and subjective techniques. Objective measures extracted from natural conversations could be more ecologically relevant, and administered more frequently – perhaps giving them added sensitivity to change. While several studies have used automated analysis methods to study autistic speech, they require manual transcriptions. In order to bypass this time-consuming process, an automated speaker diarization algorithm must first be applied. In this paper, we are testing whether a speaker diarization algorithm can be applied to natural conversations between autistic individuals and their conversational partner in a natural setting at home over the course of a clinical trial. We calculated the average duration that a participant would speak for within their turn. We found a significant correlation between this feature and the Vineland Adaptive Behaviour Scales (VABS) expressive communication score (r=0.51, p=7 × 10^-5^). Our results show that natural conversations can be used to obtain measures of talkativeness, and that this measure can be derived automatically, thus showing the promise of objectively evaluating communication challenges in ASD.

**Index Terms:** speaker diarization, autism spectrum disorder, talkativeness, natural conversations, mean length of utterance

## Introduction

Autism Spectrum Disorder (ASD) is a neurodevelopmental condition characterised by persistent deficits in social communication and interaction, the presence of repetitive and stereotyped patterns of behaviour, and sensory atypicalities^1^. These symptoms can affect job independence and the ability to form friendships, thus negatively impacting a person’s quality of life^2^. Novel therapies are under development to treat such deficits in ASD (e.g. NCT03682978; NCT04299464), however the ability to show a response is dependent on a sensitive and reliable measure of treatment effect. Treatment effects are typically assessed using scales or questionnaires that measure core symptoms and adaptive functions, which can be reported either by a patient, caregiver or clinician (e.g. Vineland Adaptive Behaviour Scales [VABS]; Burger-Caplan, Saulnier, and Sparrow 2018. Autism Behavior Inventory [ABI]; Bangerter et al. 2017). However, these assessments are time-consuming to administer, introduce elements of subjectivity (i.e. they are susceptible to placebo effects and only reflect behaviours observed by the caregiver), do not provide real-time objective information on a person’s current clinical state (and change thereof), are only infrequently administered, and do not map onto likely biological mechanisms that underpin treatment response^5–8^. Therefore, there is a critical unmet need for developing reliable and objective clinical trial endpoints that measure social communication abilities in ASD in a remote setting.

Expressive Language Sampling (ELS) through structured conversations has emerged as a promising way to measure expressive communication abilities. ELS is typically performed in a clinical setting with a trained conversation partner or physician. Several studies have utilised automated linguistic and acoustic analysis methods to analyse conversational speech to produce objective measures of speech deficits, such as the mean length of utterance (MLU) in morphemes, the number of different word roots, communication-units (C-units) per minute, and the unintelligible proportion and repetition proportion^9–12^. However, these approaches still require manual transcripts by a trained professional. In addition, most of these analyses are performed on standard clinical assessments, which are performed infrequently and in a clinical setting. In order to track changes continuously over time, more frequent speech samples are required, and ideally in a natural environment such as the participant’s home^12^. Alternative machine learning approaches that are time- and cost-efficient, and capable of analysing feature-rich datasets, have yet to be developed and deployed successfully in the context of assessing speech in individuals with ASD.

As a first step, this requires parsing conversations into segments in which the participant or conversational partner is talking. This process is known as speaker diarization, and has been widely researched with applications in conversation segmentation during meetings, phone conversations, voice verification, speech recognition etc.^13^. Speaker diarization methods typically iterate through an audio recording via a sliding window, identify the speaker(s) in each segment, and aggregate segments based on whether the same or different speaker is present. The process results in hypothesised segments of varying lengths belonging to each speaker. With the advent of deep learning, speaker diarization has become more successful in recent years^14, 15^, in part due to extremely large and publicly available training data (e.g. VoxCeleb ^16^).

However, while speaker diarization can automatically parse a conversation into a set of hypothesised speakers, it cannot know which one is the ASD participant, and which is the conversational partner. This requires the additional step of performing speaker identification, which compares the utterances from a diarized conversation with a known example of speech from the participant in question.

the motivation for this study was to assess whether or not it is possible to perform automatic speaker diarization on free-flowing conversations between an ASD participant and a conversation partner in a natural environment at home. To our knowledge, no such dataset has been collected before, and it was unknown whether an open-source pretrained speaker diarization algorithm would work in such a challenging environment. In addition, we hoped to find simple conversational features that could correlate with known clinical scales. Future work will require more advanced features using ASR and NLP to more accurately track any potential treatment effects over time, but this study has provided a proof-of-concept that such an approach could work in future clinical trials.

The goal of this study was to ascertain whether automatic speaker diarization algorithms could succeed in segmenting free-flowing conversations between ASD participants and a conversational partner in a natural environment at home. To do so, we recorded over 500 conversations from 54 ASD and 18 neuro-typical control (NTC) participants. The participants spanned in age from 5 to 45 years old. To automatically segment the conversations, we applied the open-source speaker diarization algorithm Pyannote^17, 18^. To validate its effectiveness, we manually diarized a subset of conversations. In addition to diarizing conversations, we assessed whether a simple conversation feature (the mean length of each utterance) would correlate with caregiver-reported expressive communication skills. If successful, this would lay the ground-work for future studies where more advanced methods could be applied such as automatic speech recognition (ASR) and natural language processing (NLP) to extract more complex and fine-grained conversational features that could better assess and track treatment effects over time.

## Methods

### Participant demographics

We obtained recordings from 54 ASD and 18 neurotypical control (NTC) participants, including children (5-12 years) adolescents (13-17 years), and adults (18-45 years) as part of an observational clinical trial (NCT03611075, 02/08/2018). We wanted a full span of ages to ensure that the diarization algorithm worked on children as well as adults, because most of the training data available for performing speaker diarization and speaker identification is from adult speech. The study was conducted according to the guidelines of the Declaration of Helsinki. The study has received all necessary Institutional Review Board (IRB) approvals. For the inclusion and exclusion criteria for the clinical trial, please see the Supplementary Information.

Each participant was required to have a single study partner with whom they would record their conversations. The study partners were either a staff member or family member of the participant, who must spend a few hours each day with the participant. For the inclusion criteria for the study partner, please see the Supplementary Information.

The ASD participants had their IQ measured according to the Abbreviated Battery IQ (ABIQ) of the Stanford-Binet intelligence scales fifth edition (SB5)^20^. This contains a verbal and non-verbal component, which provides an estimate of the full-scale IQ. ASD participants were subsequently placed into two categories, depending on whether their IQ was above or below 70.

While 135 participants were initially recruited into the trial, only 72 completed the conversation task at least once. See Table 1 for a break-down of the participant demographics, showing the sex of the participants (1st row: male / female), the total number of participants with recorded conversations in the study (2nd row), and the number of participants with hand-labelled recordings (3rd row). The number of conversations recorded for each cohort is displayed in brackets. Additional demographic information is shown in Supplementary Table 1.

**Table 1:** Participant Demographics. The number of participants and the number of conversations (in brackets) for each cohort. Sex (M=male, F=female). Recorded = the total number of participants (conversations) with recorded audio. Labelled = the number of participants (conversations) with hand-labelled audio. ASD = autism spectrum disorder, NTC = neuro-typical controls, Children = 5-12yr, Adolescents = 13-17yr, and Adults = 18-45yr.

|  | Children |  |  | Adolescents |  |  | Adults |  |  |  |
| --- | --- | --- | --- | --- | --- | --- | --- | --- | --- | --- |
|  | *ASD*<br>*(IQ<70)* | *ASD*<br>*(IQ>70)* | *NTC* | *ASD*<br>*(IQ<70)* | *ASD*<br>*(IQ>70)* | *NTC* | *ASD*<br>*(IQ<70)* | *ASD*<br>*(IQ>70)* | *NTC* | Total |
| **Sex (M/F)** | 6 / 1 | 12 / 2 | 4 / 4 | 6 / 0 | 7 / 3 | 5 / 3 | 8 / 1 | 5 / 3 | 0 / 2 | **53 / 19** |
| **Recorded** | 7 (49) | 14 (115) | 8 (100) | 6 (59) | 10 (78) | 8 (50) | 9 (110) | 8 (88) | 2 (3) | **72**<br>**(652)** |
| **Labelled** | 3 (4) | 7 (16) | 4 (6) | 4 (8) | 3 (3) | 3 (3) | 7 (7) | 4 (4) | 0 (0) | **35 (51)** |

### Audio recordings

We recorded natural conversations between each participant and their study partner (Figure 1*A*). The study partners were given 6 suggested topics: school, work, free-time, what they did today, dreams, and things they like or dislike. They were given no other instructions; therefore the conversations were free-flowing and unstructured in every other regard. While we did instruct them to take place in a quiet environment, they often included background noise from everyday living activities, as well as speech from other members of the household. They were instructed to be at least 5 minutes in length, but were often shorter in practice. We then restricted our analyses to those conversations lasting between 1 and 8 minutes (an empirical choice to reject accidental recordings). The audio was recorded using a smartphone with 2 microphones. The study partner was instructed to place 1 microphone facing themselves, and the other at the participant.

**Figure 1:**
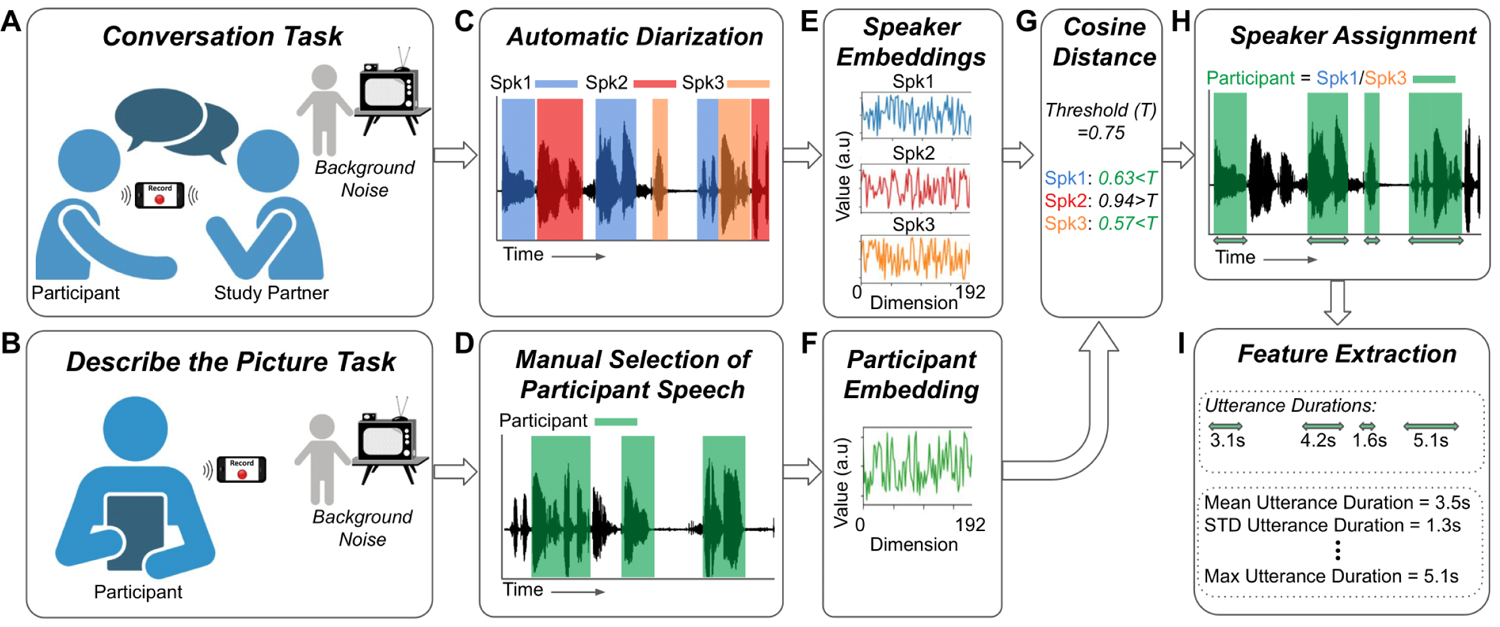
Task design, diarization methods, and feature extraction. Participants took part in **A)** a conversation with a study partner, and **(B)** a Describe the Picture (DtP) task. **C)** Conversations were automatically diarized into a hypothesised number of unknown speakers. **D)** DtP audio was manually segmented to extract participant speech only. **E and F)** The audio for each hypothesised speaker, and for the participant’s speech, were converted into fixed 192 dimensional embeddings. **G)** The distances between the speaker embeddings and the participant’s embedding were measured using the cosine distance. **H)** Speakers who’s distance was less than a predefined threshold (0.75: Spk1/Spk3 in this example) were assigned to be the participant. **I**) The duration of each utterance was measured, and their basic statistics were calculated.

In addition, we recorded a separate task in which the participants were asked to describe the contents of a picture, which we will refer to as the Describe the Picture (DtP) task (Figure 1*B*). While this task was designed to obtain different measures of ASD symptoms, it was also useful to use as an independent data set with which to obtain audio samples of the participant’s voice exclusively, so that their voice could be compared with the detected speakers in the conversation task. However, due to the presence of speech from their study partner, as well as other background talkers, we manually listened to this audio and extracted segments of the participant’s speech. We obtained between ∼30s and 60s of participant specific speech, depending on the amount available.

17 participants did not record any DtP audio. For these participants, we manually extracted their voice segments from a single conversation. We will refer to the audio from the DtP task (or the single conversation for those participants without DtP audio) as the Speaker ID audio.

### Speaker Diarization and Feature Extraction

Our speaker diarization pipeline consisted of 6 steps (Figure 1*C-I*):

1. We used the open-source speaker diarization package pyannote^17^ version 2^18^ to automatically segment each conversation into hypothesised utterances from individual speakers (Figure 1*C*). The default parameter settings were used. Given that the audio recordings were obtained in a real-world environment with potentially multiple background talkers, we did not constrain the diarization algorithm to a fixed number of speakers; instead, we allowed the algorithm to automatically estimate that number.
2. We manually extracted segments of the participant’s speech from the DtP audio recordings (Figure 1D), or from a single conversation when no DtP audio was available (which was the case for 17 participants).
3. We calculated a *speaker embedding* for the Speaker ID audio (Figure 1*F*) using the pretrained model on the hugging face platform (https://huggingface.co/speechbrain/spkrec-ecapa-voxceleb). This embedding is a fixed length 192 dimensional vector that represents any variable length speech utterance. We will refer to this embedding as the *Speaker ID embedding*. The model is based on the canonical x-vector time delay neural network (TDNN)-based architecture^14^, but with filter banks replaced with trainable SincNet features^22^.
4. By having an unconstrained number of speakers automatically detected, it’s possible that the participant’s speech could be mislabeled as 2 (or more) different speakers. To remedy this, we concatenated the audio for each hypothesised speaker together, and calculated their speaker embedding, to obtain a single embedding for each hypothesised speaker (Figure 1*E*).
5. We compared the embeddings of the hypothesised speakers with the Speaker ID embedding (from step 3) using the cosine distance. The embeddings with a distance less than a particular threshold were assigned to be the participant (Figure 1*G and H*). A threshold value of 0.75 was selected as the lowest threshold which also ensured that every participant had at least 1 conversation that was diarizable (see Supplementary Table 2). Participants can have conversations that are undiarizable if the distance between the embedding of their speaker ID audio and the embeddings of the utterances during the conversation never go below the threshold.
6. Finally, we measured the duration of each utterance belonging to the participant, and calculated their basic statistics: mean, min, max, etc. See Supplementary Table 3 for a full list.

### Diarization Performance Measurement

To test the performance of our diarization pipeline, we hand-labelled 51 conversations from 35 participants. Hand labelling consisted of listening to the conversations and labelling those segments in which either the study partner or participant was talking, or in which other environmental sounds / background talkers were occurring. We measured our ability to identify the participants speech using precision, recall, and the F1-score, which provide a percentage accuracy at identifying a target using a combination of true positives (TP), false positives (FP), and false negatives (FN):

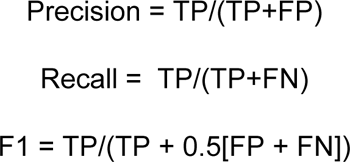

Intuitively, each of these measures can be interpreted as the following:

- Precision is a measure of how accurate the model is at correctly predicting the presence of participant speech, regardless of failed detections. For example, even if the model only predicts one instance of participant speech, and misses every other instance, as long as that single instance is correct, the ‘precision’ will be 100%.
- Recall is the opposite. It’s a measure of how accurate the model is at consistently predicting the presence of participant speech, regardless of false-positives. For example, the model could just predict that every speech utterance belongs to the participant, and this would result in a ‘recall’ score of 100%.
- The F1 score is the harmonic mean of precision and recall. The term ‘harmonic’ in this case means that it is never greater than the arithmetic mean of precision and recall, but is often smaller (i.e. the F1 score is weighted toward the smaller of precision and recall). In fact, it is equal to the arithmetic mean only when precision = recall.

### Conversation Features

From the diarization output we can extract a measure of talkativeness which is a relevant aspect of expressive communication in ASD^23, 24^. Therefore, for each conversation, we extracted the duration of each utterance for the participant (segments of speech flanked by either non-speech or speech from another talker). We will refer to this as the Utterance Duration (UD) feature. From this, we calculated basic statistics such as the mean, standard deviation, median, max, etc. (see Supplementary Table 3 for a full list) to obtain a fixed set of features for each conversation. Finally, for each statistic, we took the average across all conversations to obtain a single value per participant. We found that the majority of these statistics provided similar results (Supplementary Table 3), so for the remainder of this paper we will use the mean UD as our feature of interest. In addition, we found no relationship between the length of a conversation, and the mean UD feature (Supplemental Figure 5).

The phone used for recording the conversations had 2 microphones. The above steps were done separately for both, and then all subsequent results are taken as the average of both microphones, i.e., the features were calculated for the 2 microphones separately, and then averaged together. We did not average together correlation values or statistical results.

### Vineland Adaptive Behaviour Scales

The Vineland Adaptive Behaviour Scales (VABS^3, 25^) are commonly used for assessing various adaptive behaviours ASD. The VABS-II interview was completed for each ASD participant in this study. The communication portion of the VABS contains the subdomains of expressive and receptive communication. Expressive communication refers to the ability to express wants and needs through verbal and nonverbal communication. It is the ability to put thoughts into words and sentences in a way that makes sense and is grammatically correct. Receptive communication refers to the ability to understand and comprehend spoken language that is heard or read. Therefore, since we are using the mean UD as a conversation feature in our study, we are primarily interested in expressive communication, and will report results with regards to this clinical scale (although see Supplementary Figure 1 and Supplementary Table 3). In order to account for age differences, we used the v-scale values as opposed to the raw scores^25^. The VABS was administered at the start of the study, and 3 months later at the end. To get the most representative estimate of their communication abilities during this period, we took the average of their 2 scores. 4 participants only completed one assessment, so we used their single score.

### Statistical Analysis

All statistical analyses on group differences were performed using 2-sided Mann-Whitney *U* (MWU) tests. One-sample comparisons were performed using Wilcoxon Signed-Rank (WSR) tests. All correlations were calculated using Pearson’s correlation. The intraclass correlation coefficients are two-way random, single measures, absolute agreement: ICC(2,1).

## Results

### Diarization Performance

Figure 2 shows the performance of the diarization algorithm (precision, recall, and F1 score) at identifying the participants from the conversation recordings, split into the different cohorts (ASD [IQ<70 and IQ>70] and NTC). The precision and F1 scores were lower for IQ<70 compared with IQ>70 ASD participants (MWU: p=0.05 and 0.02, respectively), but not compared with NTC participants (p>0.05). However, no significant differences remained after correcting for multiple comparisons (Bonferroni-Holm, alpha=0.05). There were also no significant differences in recall for any cohort. The median performance across all cohorts was 76.5%, 80% and 76.5% for precision, recall and F1 Score, respectively.

**Figure 2:**
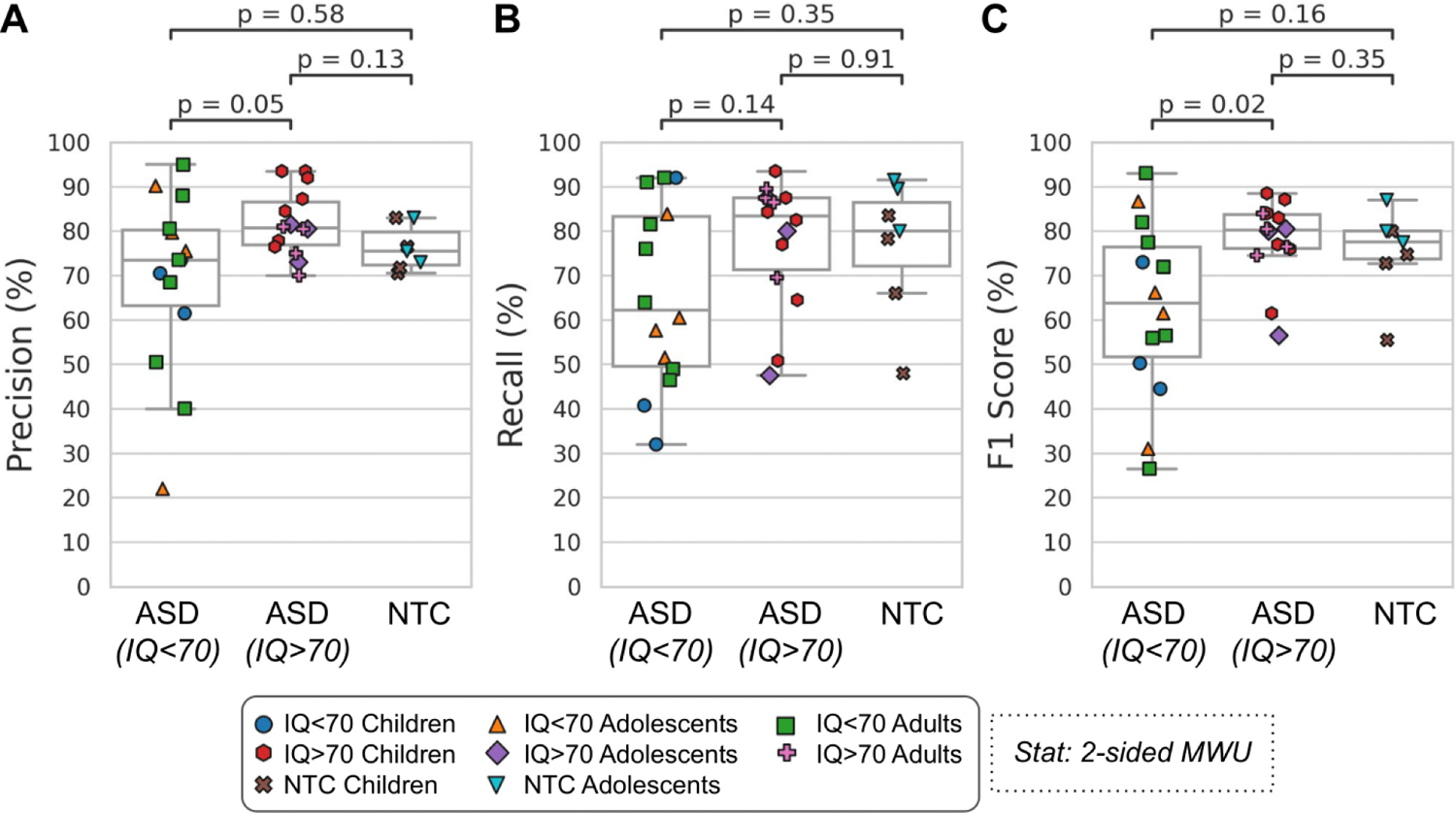
Diarization Performance. The accuracy at identifying the participant from the conversation recordings using 3 different metrics: precision **(A)**, recall **(B)** and F1 Score **(C**; see Methods). Each dot is a participant (35 participants from 51 conversations). ASD = autism spectrum disorder, NTC = neurotypical control. Statistics are calculated using a 2-sided Mann-Whitney U test.

### Correlation between True and Predicted Conversation Features

Figure 2*A* shows a scatter plot and correlation between the true and predicted mean UD conversation feature (r=0.81, p=3×10^-9^). Figure 2*B* shows a boxplot illustrating the difference between the true and predicted mean UD for each cohort. There was no significant difference between the different cohorts (MWU: p > 0.05). The ASD (IQ<70) cohort was significantly greater than zero (WSR: p=0.02).

### Comparing ASD and NTC Conversation Features

Having validated that our speaker diarization method could identify the participants from the hand-labelled conversations (35 participants from 51 conversations) and predict the mean UD conversation feature, we then extended our analyses to the full dataset of participants (72 participants from 652 conversations). We then compared the conversation features between each cohort, after calculating the mean value for each participant. Figure 4 shows a boxplot illustrating the differences in the mean UD between the cohorts, when using the true labels (**A**; 35 participants from 51 conversations) and the predicted labels (**B**; 72 participants from 652 conversations). *Note that due to the different number of conversations used to calculate the mean value for each participant, the values reported in **A** and **B** are not directly comparable.* In both cases, ASD (IQ<70) participants had significantly shorter Mean Response Durations than ASD (IQ>70) and NTC participants (p < 0.05). There were no significant differences between ASD (IQ>70) and NTC participants.

After correcting for multiple comparisons (Bonferoni-Holm; n=3, alpha=0.05), there was no longer a significant difference between the ASD (IQ<70) and NTC cohorts for the Predicted Labels (p-corrected = 0.07024).

### Correlation with VABS Communication Scores

While categorising ASD participants based on their IQ gives an indication of communication challenges, the VABS expressive communication (EC) score gives a more appropriate clinical measure of communication abilities. Figure 5 shows a scatter plot between the mean UD (x-axis; logarithmic scale) and the VABS EC score (y-axis) for the True labels **(A)** and the Predicted labels **(B)**. In both cases, there was a significant correlation (r=0.5, p=0.007, and r=0.51, p=7×10^-5^, respectively).

Given how we observed a significant difference in mean UD between the ASD (IQ<70) and ASD (IQ>70) groups (Figure 4), we wondered if it was primarily IQ that was driving the correlations with the VABS EC score. Indeed, IQ and VABS EC are correlated with each other (r=0.39, p=0.004). To take into account this possibility, we performed a partial correlation between the mean UD and the VABS EC score, using the IQ as a covariate. Doing so still resulted in a significant correlation (r=0.43, CI95% = [0.18, 0.63], p=0.001).

We also calculated the correlations for each age group separately for the predicted mean UD feature (Supplementary Figure 4). In spite of the smaller sample sizes, we still found significant correlations for the children (Pearson’s r = 0.44, p = 0.04) and adult groups (r = 0.68, p = 0.002), and a trend towards a significant correlation for the Adolescent group (r = 0.44, p = 0.09).

### Test-Retest Reliability

We calculated the intraclass correlation coefficient (ICC(2,1); two-way random, single measures, absolute agreement) of the predicted mean UD feature to measure its consistency across conversations (r = 0.46). Due to the fact that we only had ∼1 conversation with hand-labels per participant, we could not calculate an ICC value for the true labels.

## Discussion

Using an automatic speaker diarization algorithm we were able to use natural conversations to identify ASD participants as they conversed with a study partner. The algorithm could quite accurately capture a conversation feature related to talkativeness which we refer to as the mean Utterance Duration (UD): the average length of time (in seconds) that a participant would speak for within their turn (Figure 3). This feature was significantly different between ASD participants with an IQ<70 and those with an IQ>70 (Figure 4), but not between the ASD (IQ>70) and NTC groups. Finally we found that the mean UD feature exhibited a significant correlation with a clinical measure of communication deficits in ASD: the Vineland Adaptive Behaviour Scales (VABS) expressive communication subdomain score (Figure 5).

**Figure 3:**
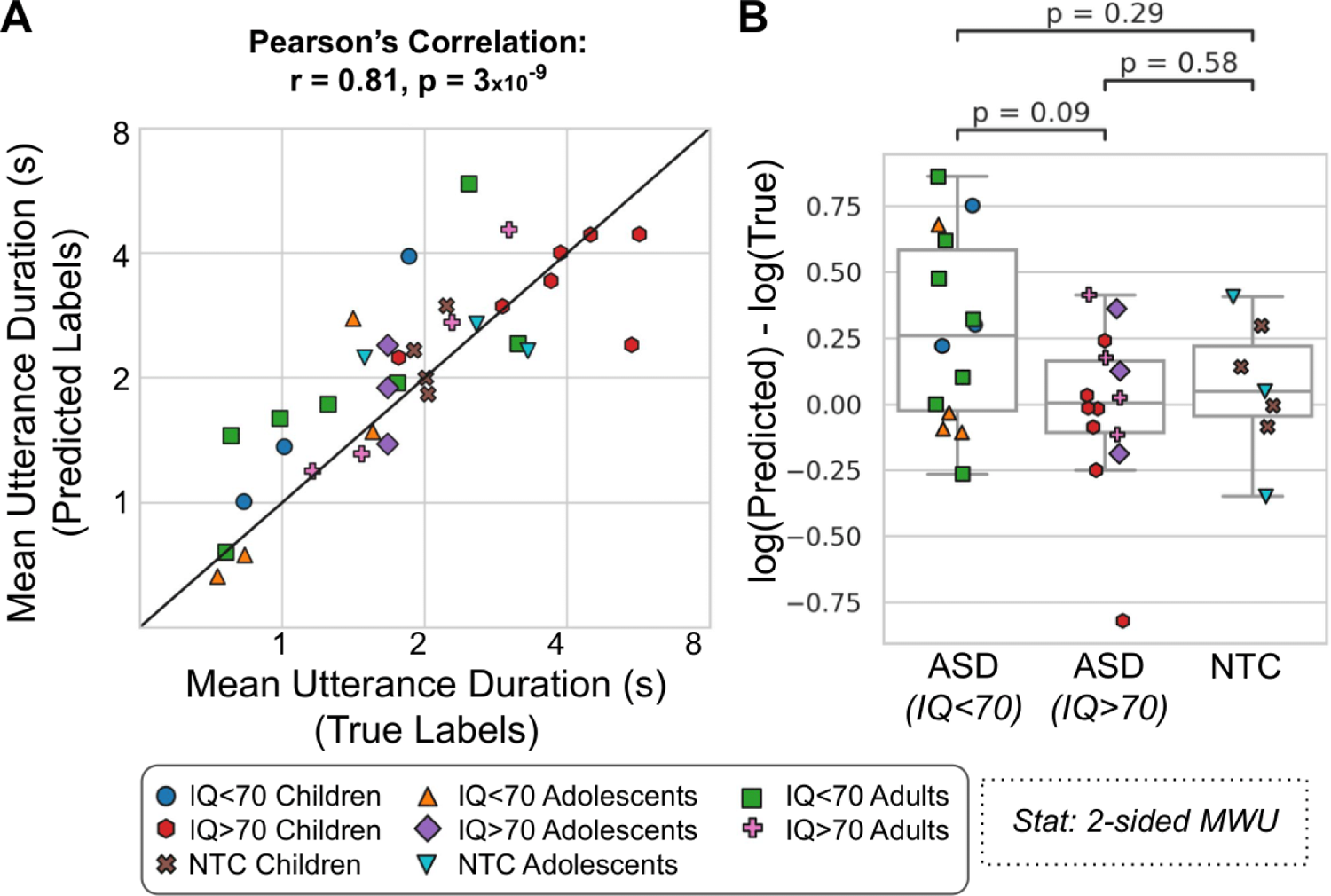
Mean Utterance Duration (UD) - Comparison of True and Predicted Labels. **A)** Pearson’s correlation between the true and predicted (x and y-axes, respectively; both on logarithmic axes) mean UDs. **B)** The difference (y-axis) between the true and predicted mean UD for each cohort (x-axis). The ASD (IQ<70) cohort was significantly greater than zero (Wilcoxon Signed-Rank test: p=0.02). Each dot is a participant (35 participants from 51 conversations). ASD = autism spectrum disorder, NTC = neurotypical control.

**Figure 4.**
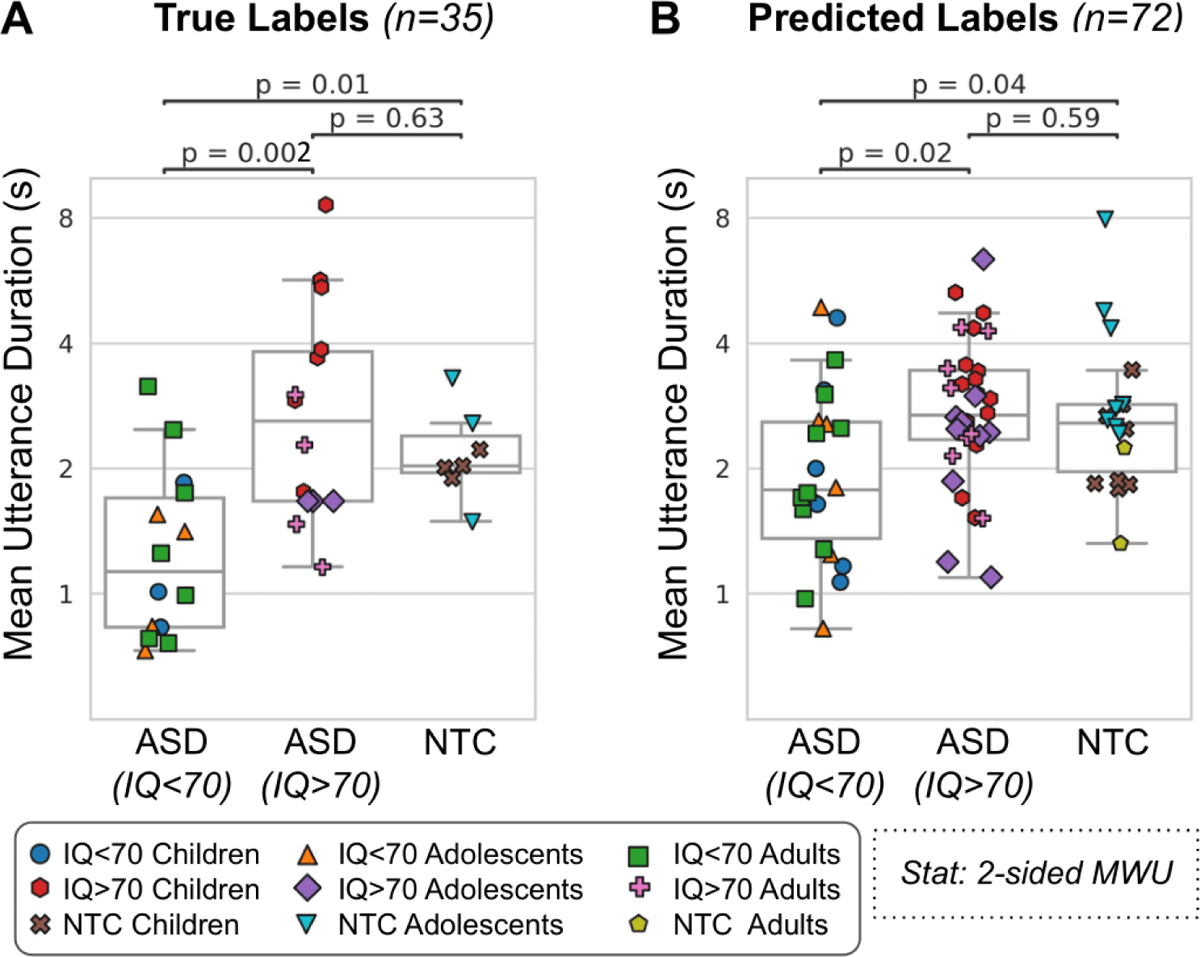
Mean Utterance Duration (UD) - Cohort Comparison. The mean UD (y-axis; logarithmic scale) for each cohort (x-axis) for the true labels **(A**; 35 participants from 51 conversations) and predicted labels **(B**; 72 participants from 652 conversations). ASD = autism spectrum disorder. NTC = neurotypical control.

**Figure 5.**
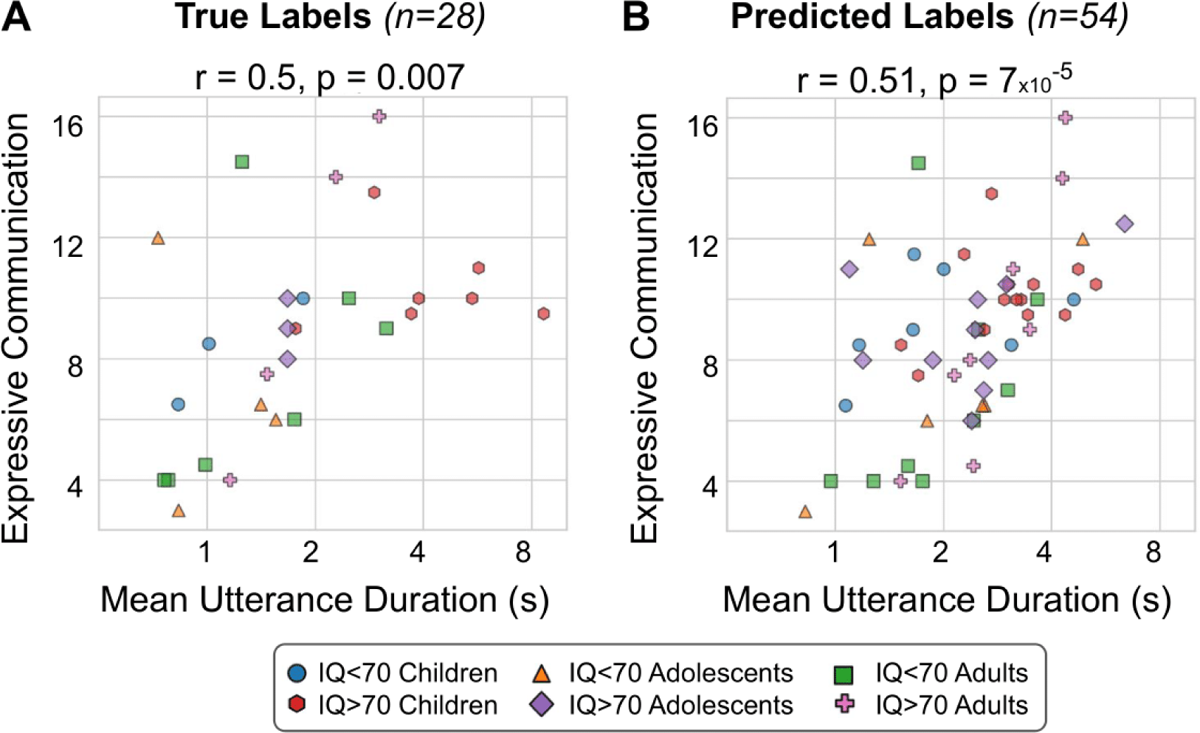
Mean Utterance Duration (UD) - Comparison with VABS. The correlation (Pearson’s r) between the mean UD (x-axis; logarithmic scale) and the VABS expressive communication score (y-axis) for the true labels **(A**; 28 participants from 42 conversations) and predicted labels **(B**; 54 participants from 499 conversations). Each dot represents an ASD participant.

Our mean UD feature should be distinguished from the Mean Length of Utterance (MLU) feature that is commonly used in ASD research^23^. The MLU is the average number of morphemes per utterance. Another measure of talkativeness explored in ASD research calculates the number of ‘Communication-units’ or ‘C-units’ per minute^26^, which requires knowledge of sentential and phrasal structure^9, 24^. Both the MLU and C-unit features require manual transcripts in order to be calculated; our mean UD feature is therefore rather simple by comparison, as it only requires the temporal labels from the diarization. They are also technically independent of the duration of the utterance in seconds, but are likely to correlate strongly with it. Further research combining manual transcripts with diarization will be required in order to do a direct comparison.

While the diarization algorithm can automatically segment a conversation into different talkers, identifying the participant’s voice requires the additional manual step of listening to and extracting examples of the participant’s voice from separate recordings. This is necessary because the diarization algorithm can’t know apriori who the participant is. While this means that the analysis can’t be fully automated, it ensures that only a minimal amount of manual labour is required, as only a minute or so of participant speech is required in order to diarize an unlimited amount of conversations. One potential method to automate this step would be for a clinician to record audio of the participant in a controlled environment. Whether or not this would work in practice, and would be cheaper / quicker than manually listening to and diarizing a short amount of audio, remains to be seen. However, recording examples of participant speech at an in-clinic setting would alleviate any privacy concerns of listening to conversations recorded at home.

In order to identify the participant’s voice from the number of hypothesised speakers from the diarization algorithm, we compared the distance of their embedding vectors using the cosine distance. We selected a fixed threshold, which was the lowest value at which every participant had at least one conversation that was diarizable (Supplementary Table 1). However, selecting a fixed threshold is probably not optimal given the heterogeneity of our data (different recording environments, different sexes/ages of the participants and their study partners, and different clinical diagnoses). One possibility is to manually diarize (at least) one conversation per participant, and then choose the threshold to optimise the differentiation between the two talkers. Alternatively one could train a model to optimally differentiate between the two predefined speakers, given that their speech would be known apriori. In addition, the speaker ID network^22^ could be fine tuned by incorporating ASD speech into its training data.

Despite having 2 microphones on our recording device, we only performed single-channel speaker diarization, and then averaged the results from the 2 microphones. The study partner was instructed to place 1 microphone facing themselves, and the other at the participant, however it was not possible to confirm if this was carried out. Future work could use more sophisticated diarization algorithms that incorporate both channels in the model training, thus resulting in an optimised dual-channel speaker diarization system^27, 28^.

The expressive communication survey measures a wide variety of communication abilities, including non-verbal communication, so we are unlikely to fully capture this clinical measure using a simple conversation feature such as the mean UD^24^. In addition, the VABS questionnaire suffers from placebo effects due to its subjective nature^8^. In future, we will look at additional low-level conversation features such as pitch variation, prosody, speech duration, speech rate, pauses, and turn-taking rate^29^, as well as high-level features extracted using automatic speech recognition (ASR) such as self-repetition or repeating the study partner (echolalia^30^), and restricted interests^31^. This would also help to explain the lack of any difference in mean UD between the ASD (IQ>70) and NTC groups (Figure 4); this is probably because the mean UD feature is too simple to detect any differences, with more complex features from ASR being required instead.

It should be noted that there are several limitations with regards assessing ASD symptoms by recording natural conversations at home. For example, using untrained conversational partners, each with their own unique communication abilities and motivational levels, will undoubtedly have an impact on the conversational style and talkativeness of the participant in question. In addition, environmental noise and background talkers can be distracting when trying to have a conversation, and could lead to reduced social interaction. Future studies will need to carefully consider the effects that these issues may have when assessing any potential drug effects.

The ASD (IQ<70) cohort had the poorest diarization performance, particularly with regards to precision (Figure 2). Low precision means that the algorithm would often mistake the study partner speech for participant speech. This led to an overestimation of the UD feature for these participants (Figure 3). However, this overestimation was not drastic enough to radically alter their resulting features, meaning that we still observed shorter UDs for these participants. One possible explanation for this discrepancy would be that the study partner’s speech mirrored the participant’s speech. Indeed, when comparing the UD features for the 2 groups, we found a small correlation (r=0.28, p=0.09, absolute difference = 0.33+/-1.7s; Supplementary Figure 2). In addition, we found a correlation between the study partner’s mean UD feature and the VABS expressive communication score (r = 0.37, p=0.06; Supplementary Figure 3). Indeed, prior research has suggested that speech and language features produced by conversational partners can be used to predict an autism diagnosis^32, 33^. In contrast, ASD participants tend not to adapt their talkativeness based on their partner’s behaviour^34^. Together, these data suggest that low diarization precision does not necessarily lead to a poor estimation of the UD of a participant. In spite of the mean UD feature being relatively unaffected by poor precision, other higher-level features obtained using automatic speech recognition would be strongly affected.To remedy this, future work will require hand-labelling dozens of conversations with ASD participants in order to fine-tune the diarization algorithm. It should also be noted that the results of our study may not be generalizable to other neurodevelopmental disabilities (e.g., Down syndrome). Further research would be needed to evaluate the efficacy of our algorithm in such contexts. However, the field of automatic speaker diarization is still evolving, and future research may develop more effective and robust algorithms for a wider range of populations and contexts.

## Data Availability

Anonymized features are available upon reasonable request. Please contact Florian Lipsmeier

## Acknowledgments

We gratefully acknowledge the individuals with ASD and their families. We also thank the input and support of Federico Bolognani, former employee of F. Hoffmann-La Roche Ltd. and the contribution of the Roche Digital Biomarker team in Pharma Research and Early Development Informatics, in particular Raphael Ullmann, Jan Beckmann, Emma Poon and Samuel Nova, for their various roles in app development, data collection during the clinical trial and input into data analysis.

## Author Contributions

JOS and GB performed the analyses with support from WC, DS, and NM. JOS wrote the manuscript with support from GB, WC, DN, and FL. CC, JH, FL, and DN, conceived the study and analysis plan. PS, JT, EE, TK, LM, JH, ML, FL, conceived and implemented the study. All authors discussed the results and contributed to the final manuscript.

## Data availability statement

Anonymized features are available upon reasonable request. Please contact Florian Lipsmeier

## Additional Information

The authors declare no Competing Non-Financial Interests but the following Competing Financial Interests: JOS, GB and JT are full-time contractors of F. Hoffmann-La Roche Ltd. WC, DS, PS, EE, TK, LM, ML, and FL are full-time employees of F. Hoffmann-La Roche Ltd. DN and JH are full-time employees and shareholders of F. Hoffmann-La Roche Ltd. CC is a full-time employee of Genentech, and a shareholder of F. Hoffmann-La Roche Ltd. NM does not possess any competing interests.

## Institutional Review Board Statement

Informed consent was obtained from all subjects and/or their legal guardian(s) (incase of minors). The study was conducted according to the guidelines of the Declaration of Helsinki, and approved by Advarra IRB (Pro00035060, May 2018), BRANY IRB (#18-10-193-01, August 2018), Nathan Kline Institute for Psychiatric Research/Rockland Psychiatric Center IRB (1218792-12, August 2018), the Holland Bloorview Research Ethics Board (18-800, November 2018), the Wales Research Ethics Committee 5 Bangor (251572, September 2018), and the Western Institutional Review Board (20191384, August 2019).

## Supplementary

### Inclusion and Exclusion Criteria

The inclusion criteria for participants with ASD were as follows:

- Diagnosis of ASD based on the Diagnostic and Statistical Manual of Mental Disorders (DSM-5), and the Autism Diagnostic Observation Schedule (ADOS-2).
- Children’s Yale-Brown Obsessive Compulsive Scale modified for ASD (CY-BOCS-ASD) total score of at least 12.
- Clinical Global Impression-Severity (CGI-S) score of at least 4 about participant’s current autism severity.
- Intelligence quotient (IQ) score of 50 or above as assessed by the Abbreviated Intelligence Quotient (ABIQ) SB5 scale.
- English proficiency compatible with the study measurements as judged by the investigator.
- Hearing, vision, and speech compatible with the study measurements as judged by the Investigator.
- All medications and treatments were expected to be stable for the duration of the study.

The diagnostic evaluations were completed at the study site by research staff and supervised by a licensed psychologist.

The exclusion criteria for all participants were as follows:

- Participation in an investigational drug or device study within 4 weeks, or five times the half-life (if it is a drug study) of the investigational molecule (whichever is longer), prior to screening and the participant is expected not to enrol in any other trial during the study.
- Co-occurring disease or condition that could interfere with, or treatment which might interfere with, the conduct of the study, or that would, in the opinion of the Investigator, pose an unacceptable risk to the participant in this study.
- Unstable or uncontrolled clinically significant psychiatric and/ or neurological disorder that may interfere with the objectives of the study.
- Participants with known “syndromic” ASD (e.g., Fragile-X syndrome, Angelman Syndrome, Prader-Willi, Rett’s syndrome, tuberous sclerosis, Dup15q syndrome).
- History of alcohol misuse and/ or illicit drug use during the last 12 months prior to screening.

The inclusion criteria for the study partners were as follows:

- A staff member of the residential home can be the caregiver if this person spends sufficient time with the participant. In the opinion of the Investigator, the caregiver must be able to reliably assess the participant’s mental status, activities, and behavior, and report on the participant’s adherence and health. This would normally be possible when the caregiver spends a few hours each day with the participant.
- A family member living at the participant’s home can be the caregiver if the participant returns home every night. When the participant returns home only over the weekend, a family member can only be the caregiver if they have intensive interaction with the participant during the week e.g., via phone calls, calls via Skype, SMS messages, etc. The quality of these interactions between caregiver and participant needs to be assessed for each participant to determine whether they are sufficient.

**Supplementary Table 1:** Participant Demographics Additional demographic information showing the sex, age, and IQ of each cohort. ASD = autism spectrum disorder. NTC = neurotypical control. In addition, the ethnicity breakdown across all cohorts is as follows: 72.2% white, 7% Asian, 18% black or African American, and 2.7% multiple.

| Cohort | Sex | Count | Age |  | IQ |  |
| --- | --- | --- | --- | --- | --- | --- |
|  |  |  | *Mean* | *STD* | *Mean* | *STD* |
| ASD (IQ<70) | Female | 2 | 12.0 | 8.5 | 57.0 | 9.9 |
|  | Male | 20 | 16.7 | 6.9 | 59.3 | 6.6 |
| ASD (IQ>70) | Female | 8 | 17.8 | 6.9 | 80.9 | 8.5 |
|  | Male | 24 | 15.0 | 8.1 | 91.9 | 13.9 |
| NTC | Female | 9 | 17.2 | 12.8 | 100.7 | 12.5 |
|  | Male | 7 | 11.7 | 4.2 | 97.4 | 10.9 |

**Supplementary Table 2:** Cosine Distance Threshold Selection Threshold: The threshold (left column) for the cosine distance metric between the participant’s Speaker ID embedding and the embeddings of the speakers automatically identified by the diarization algorithm (see Figure 1). N participants: The number of participants successfully diarized (out of 35). Participants can have conversations that are undiarizable if the distance between the embedding of their speaker ID audio and the embeddings of the utterances during the conversation never go below the threshold. F1 Score: The accuracy at identifying the participant from the conversation recordings (averaged across all participants; see Figure 2). Pearson’s r,p: The correlation between the true and predicted mean UD (see Figure 3).

| Threshold | N participants | F1 Score | Pearson's r,p |
| --- | --- | --- | --- |
| 0.50 | 33 | 0.70 | $r=0.82, p=6 \times 10^{-9}$ |
| 0.55 | 33 | 0.72 | $r=0.82, p=7 \times 10^{-9}$ |
| 0.60 | 34 | 0.72 | $r=0.81, p=5 \times 10^{-9}$ |
| 0.65 | 34 | 0.72 | $r=0.81, p=5 \times 10^{-9}$ |
| 0.70 | 34 | 0.72 | $r=0.81, p=4 \times 10^{-9}$ |
| 0.75 | **35** | 0.71 | $r=0.81, p=3 \times 10^{-9}$ |
| 0.80 | **35** | 0.70 | $r=0.79, p=2 \times 10^{-8}$ |
| 0.85 | **35** | 0.69 | $r=0.79, p=2 \times 10^{-8}$ |

**Supplementary Table 3:**
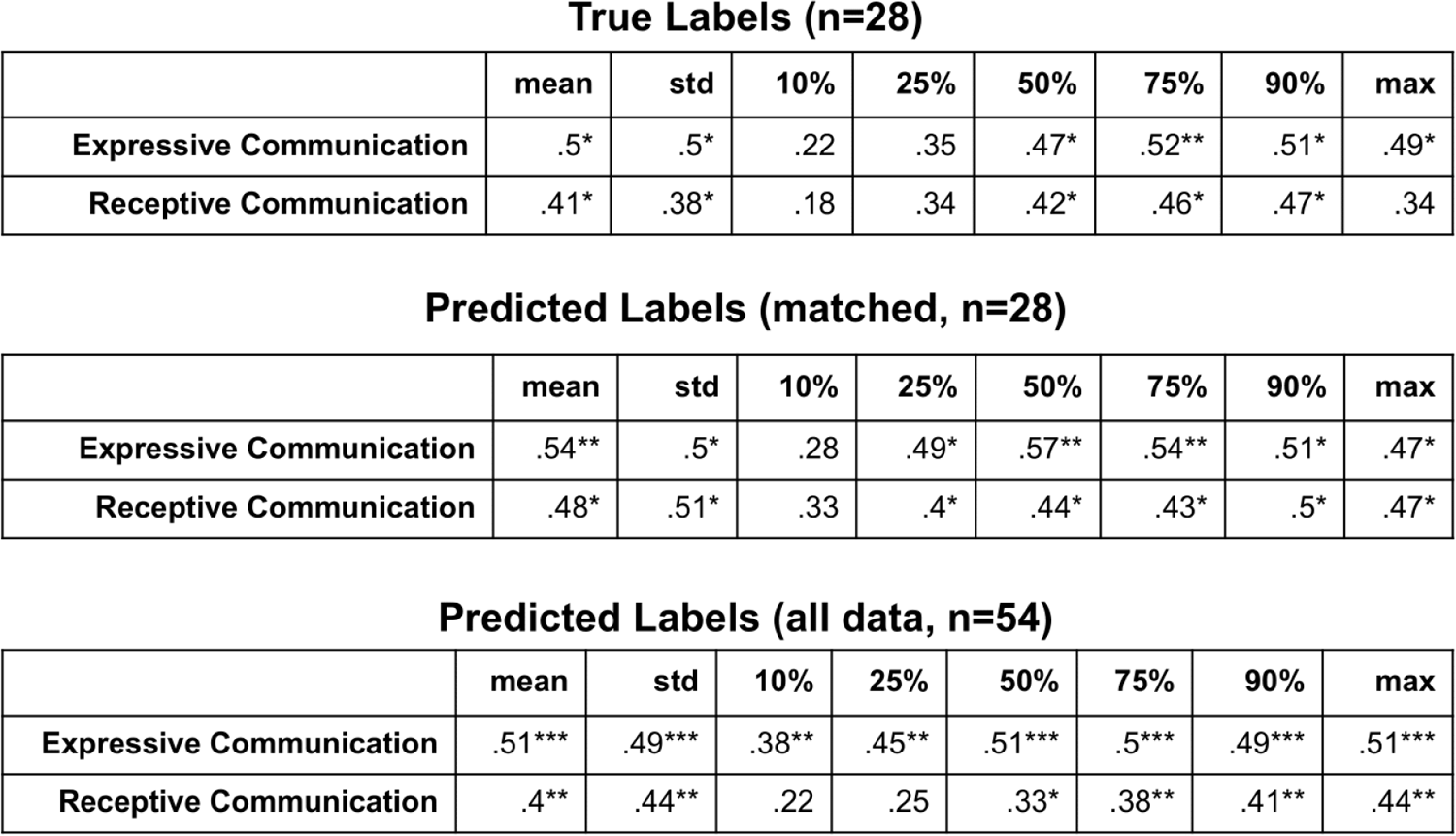
Utterance Duration (UD) statistics and their correlation with the VABS communication scales. The UD can be summarised using a variety of statistics. For the main paper, we showed the results using the mean UD. Here, we are showing the results from all statistics (columns) and their correlation with the VABS expressive and receptive communication scores (rows). In addition, we report the correlations for the predicted labels when using the same subset of conversations as the true labels (middle; Predicted Labels (matched, n=28)).

True Labels (n=28)
|  | mean | std | 10% | 25% | 50% | 75% | 90% | max |
| --- | --- | --- | --- | --- | --- | --- | --- | --- |
| **Expressive Communication** | .5* | .5* | .22 | .35 | .47* | .52** | .51* | .49* |
| **Receptive Communication** | .41* | .38* | .18 | .34 | .42* | .46* | .47* | .34 |

Predicted Labels (matched, n=28)
|  | mean | std | 10% | 25% | 50% | 75% | 90% | max |
| --- | --- | --- | --- | --- | --- | --- | --- | --- |
| **Expressive Communication** | .54** | .5* | .28 | .49* | .57** | .54** | .51* | .47* |
| **Receptive Communication** | .48* | .51* | .33 | .4* | .44* | .43* | .5* | .47* |

Supplementary Table 3: Utterance Duration (UD) statistics and their correlation with the VABS communication scales.
|  | mean | std | 10% | 25% | 50% | 75% | 90% | max |
| --- | --- | --- | --- | --- | --- | --- | --- | --- |
| **Expressive Communication** | .51*** | .49*** | .38** | .45** | .51*** | .5*** | .49*** | .51*** |
| **Receptive Communication** | .4** | .44** | .22 | .25 | .33* | .38** | .41** | .44** |

**Supplementary Figure 1:**
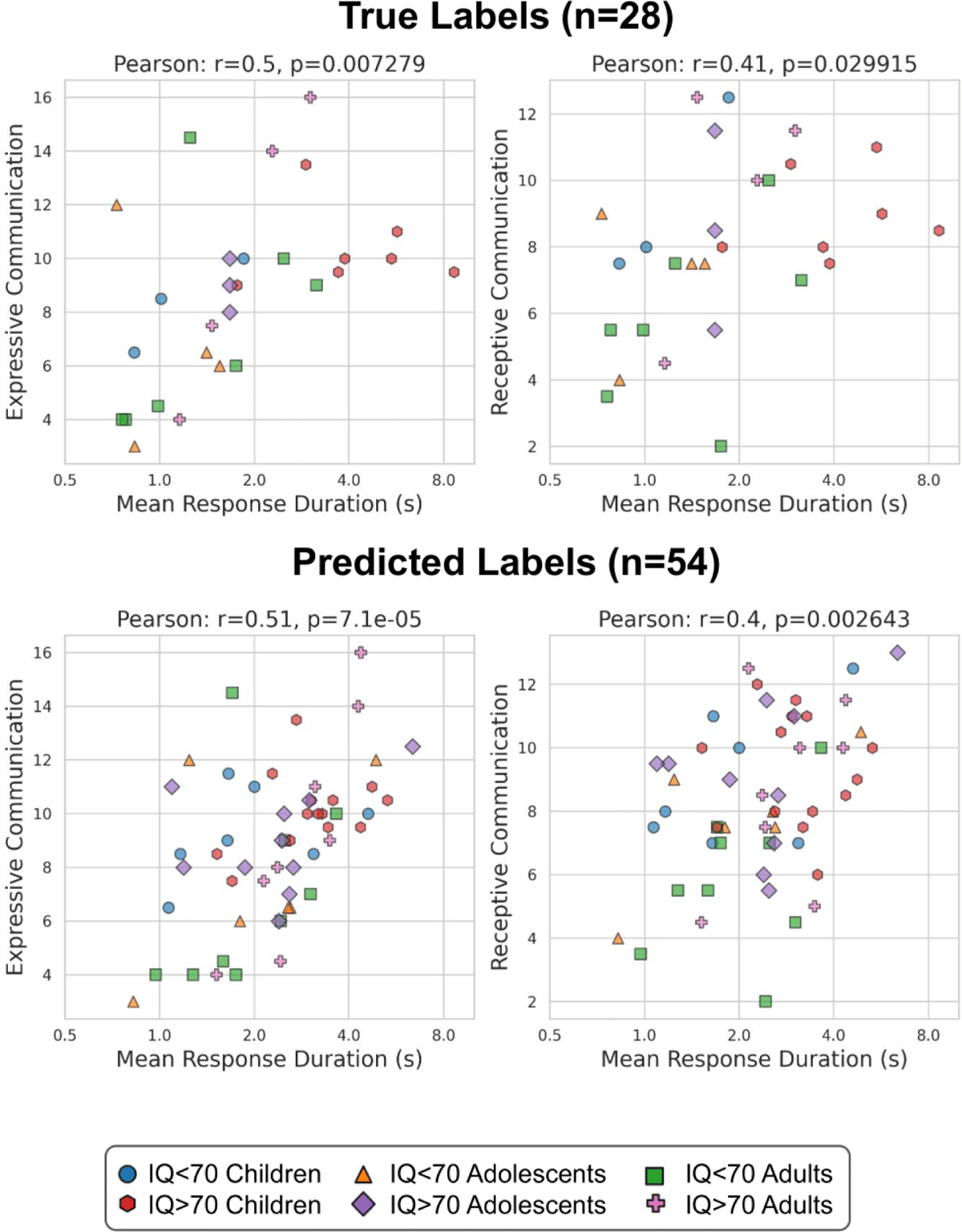
Mean Utterance Duration (UD) - Comparison with VABS. The correlation (Pearson’s r) between the mean UD (x-axis; logarithmic scale) and the VABS expressive communication score (y-axis; left column) and the receptive communication score (right column) for the true labels (top row: 28 participants from 42 conversations) and predicted labels (bottom row: 54 participants from 499 conversations). Each dot represents an ASD participant.

**Supplementary Figure 2:**
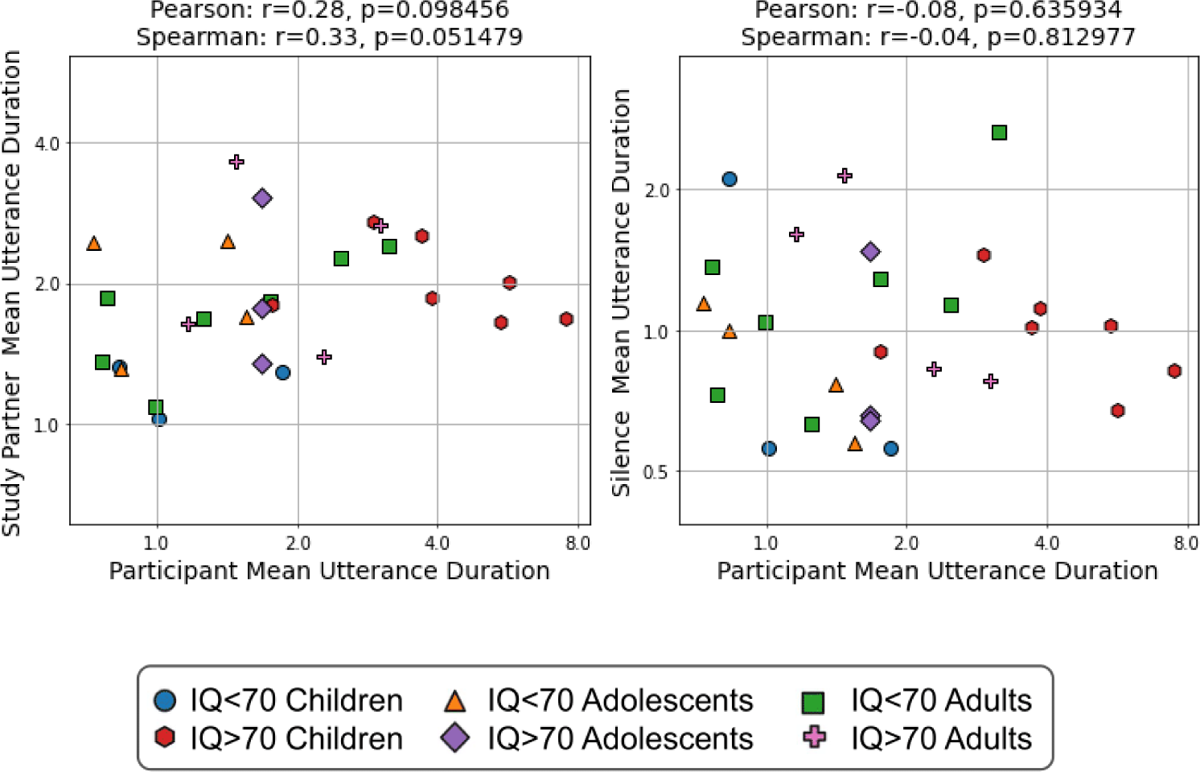
Mean Utterance Duration (UD) - Participant and Study Partner Comparison. The correlation between the Participant’s and Study Partner’s mean UD (left) and between the Participant’s and Silence mean UD (right) using the hand-labelled data. Each dot represents an ASD participant.

**Supplementary Figure 3:**
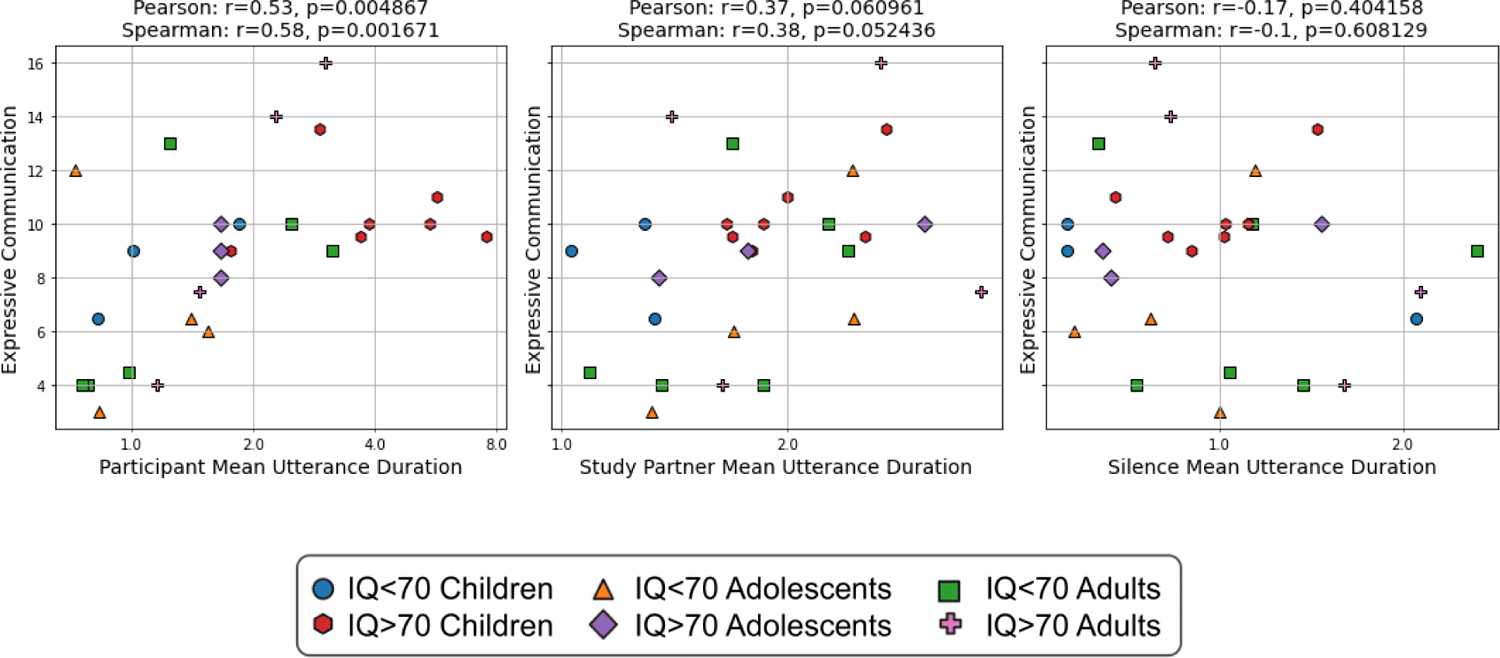
Mean Utterance Duration (UD) - Comparison with VABS. The correlation (Pearson’s r) between the mean UD (x-axis; logarithmic scale) and the VABS expressive communication score (y-axis) for the participant’s speech (left), study partner speech (middle) and silence (right) using the hand-labelled data. Each dot represents an ASD participant.

**Supplementary Figure 4:**
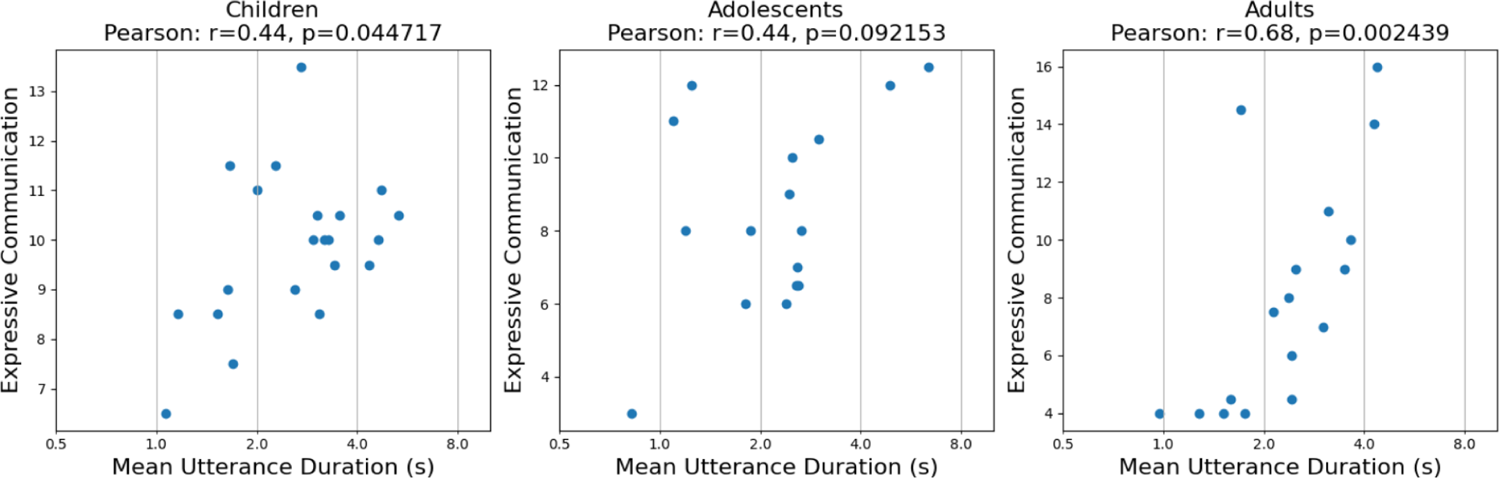
Mean Utterance Duration (UD) vs. VABS - Age Comparison. The correlation (Pearson’s r) between the mean UD (x-axis; logarithmic scale) and the VABS expressive communication score (y-axis) for each age group (left: children, middle: adolescents, right: adults). Each dot represents an ASD participant.

**Supplementary Figure 5:**
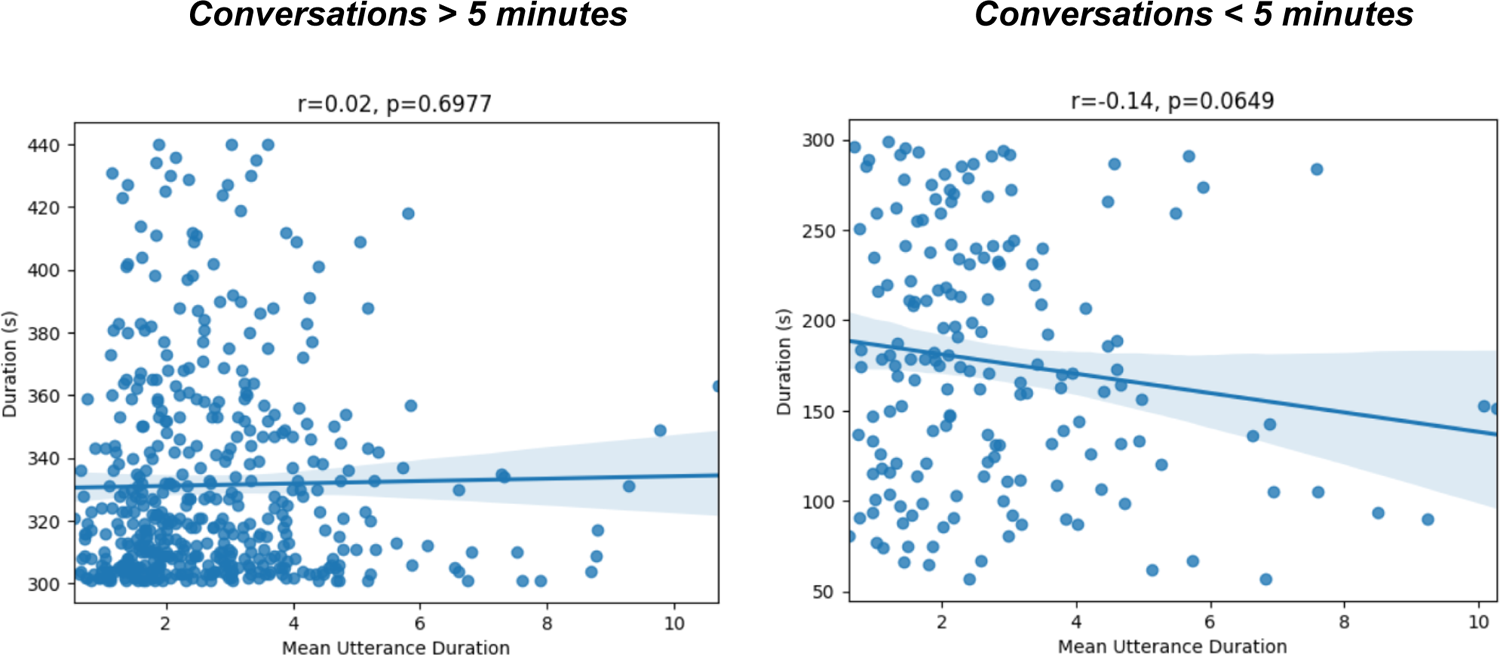
Mean Utterance Duration (UD) vs. Conversation Length. The correlation (Pearson’s r) between the mean UD (x-axis) and the conversation duration (s; y-axis) for conversations that were greater than 5 minutes (left) and less than 5 minutes (right). Each dot represents a conversation.

